# Predictive Structured-Unstructured Interactions in EHR Models: A Case Study of Suicide Prediction

**DOI:** 10.1101/2021.08.10.21261831

**Authors:** Ilkin Bayramli, Victor Castro, Yuval Barak-Corren, Emily M. Madsen, Matthew K. Nock, Jordan W. Smoller, Ben Y. Reis

## Abstract

Clinical risk prediction models powered by electronic health records (EHRs) are becoming increasingly widespread in clinical practice. With suicide-related mortality rates rising in recent years, it is becoming increasingly urgent to understand, predict, and prevent suicidal behavior. Here, we compare the predictive value of structured and unstructured EHR data for predicting suicide risk. We find that Naive Bayes Classifier (NBC) and Random Forest (RF) models trained on structured EHR data perform better than those based on unstructured EHR data. An NBC model trained on both structured and unstructured data yields similar performance (AUC = 0.743) to an NBC model trained on structured data alone (0.742, *p* = 0.668), while an RF model trained on both data types yields significantly better results (AUC = 0.903) than an RF model trained on structured data alone (0.887, *p*<0.001), likely due to the RF model’s ability to capture interactions between the two data types. To investigate these interactions, we propose and implement a general framework for identifying specific structured-unstructured feature pairs whose interactions differ between case and non-case cohorts, and thus have the potential to improve predictive performance and increase understanding of clinical risk. We find that such feature pairs tend to capture heterogeneous pairs of general concepts, rather than homogeneous pairs of specific concepts. These findings and this framework can be used to improve current and future EHR-based clinical modeling efforts.

## Introduction

In recent years there has been a proliferation of clinical prediction models powered by electronic health records (EHRs). Many prediction models rely primarily on structured data from the EHR, which typically includes diagnostic, laboratory, medication, and procedure codes. Yet most EHRs also contain unstructured data such as clinician notes, which may include information already captured in the structured data, as well as information not present in the structured data (Figure 1). Unstructured EHR data have been used for clinical predictive tasks, both as a standalone feature-set and in combination with structured data. ^1,2,3,4^

**Figure 1.**
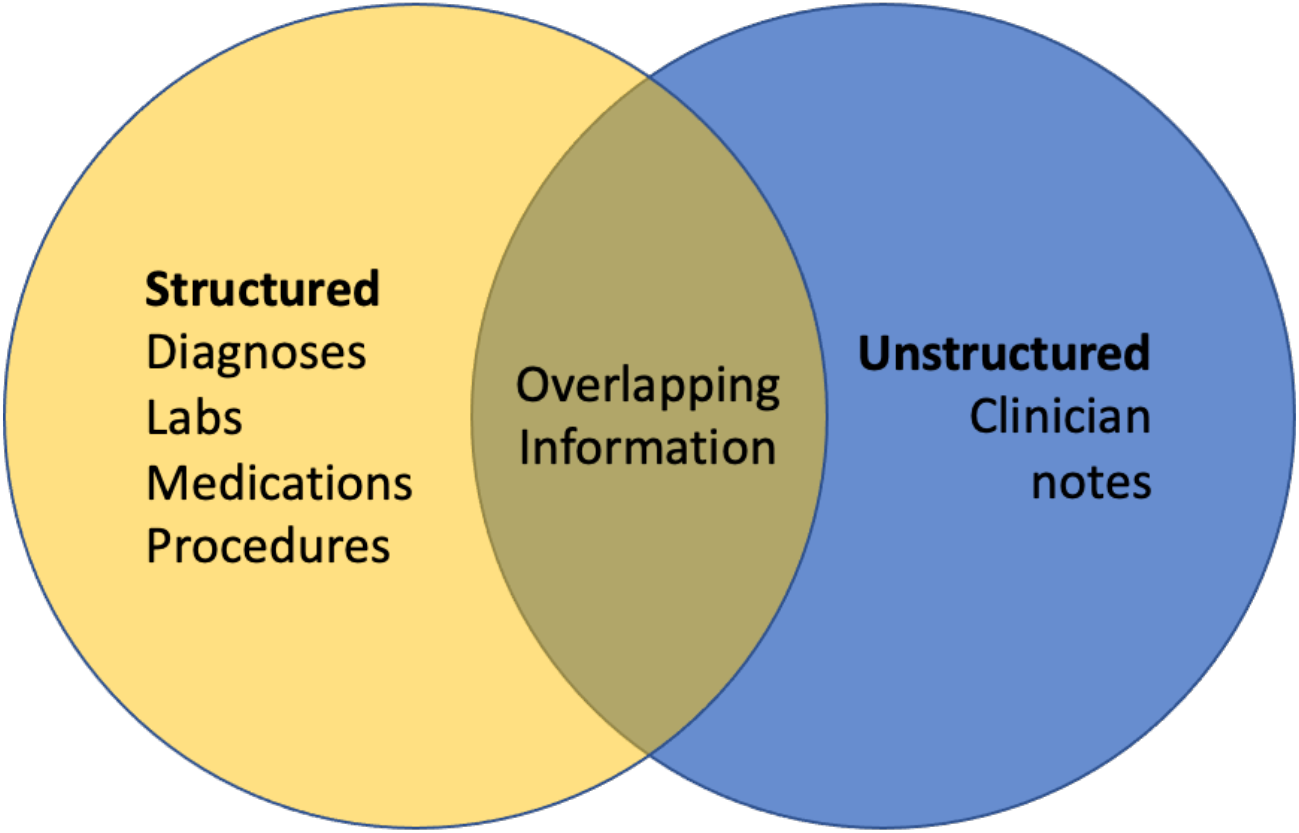
Information overlap in EHR data. Electronic health records contain both structured and unstructured data. These two types of data contain both unique and overlapping information.

In order to optimally integrate both structured and unstructured data and improve predictive performance, it is important to understand the predictive value of each data type. It is also important to understand the interactions between these two data types and identify instances where the nature of these interactions differs between case and non-case populations. Such differences can be valuable for deepening our understanding of clinical risk and for improving clinical risk prediction in models that are able to capture these interactions.

As a case study, we focus on suicide prediction. Approximately 800,000 people die by suicide every year worldwide, accounting for 1.5% of all deaths.^5^ Suicide is the tenth leading cause of death in North America and a leading cause of death globally among persons 15 to 24 years of age.^6^ With suicide-related mortality rates rising in recent years,^7^ it is becoming increasingly urgent to understand, predict, and prevent suicidal behavior. Early and accurate identification of individuals with elevated risk for suicide attempts is critical for developing effective suicide prevention strategies. Predicting suicide risk, however, is a complex challenge. The intuition of clinicians for detecting at-risk individuals is no better than random chance,^8^ underscoring the potential value of algorithmic approaches to this challenge.

In recent years, rapidly growing quantities of electronic health data along with advancements in statistical learning methods have enabled the development of suicide risk prediction models. We recently developed one such model using data from over 1.7 million patients in a large healthcare system (Mass General Brigham);^9^ the model detected 45% of suicide attempts an average of 3 to 4 years in advance, with a specificity of 90% and an area under the receiver operating curve (AUC) of 0.77. Since structured EHR data capture only some elements of clinical presentation, in the present study we seek to improve upon this prediction accuracy by examining features extracted using natural language processing (NLP) from unstructured clinician notes. (For simplicity, we refer to these as “unstructured features.”)

The goals of this study are threefold: (1) To compare the predictive value of structured and unstructured EHR data as standalone datasets for predicting suicide risk; (2) To evaluate the increase in prediction performance when integrating both structured and unstructured data using various models; and (3) To identify structured-unstructured feature pairs in which the interaction between the two features differs substantially between case and non-case populations, and which may thus have the potential to improve predictive performance. To achieve the latter, we propose a framework for identifying structured-unstructured feature pairs in which the interaction between the two features differs significantly between case and non-case cohorts.

## Methods

We analyzed data from the Mass General Brigham Research Patient Data Registry (RPDR),^10^ an EHR data warehouse covering 4.6 million patients from two large academic medical centers in Boston, MA, USA (Massachusetts General Hospital and Brigham and Women’s Hospital), as well as their affiliated community and specialty hospitals in the Boston area. The RPDR was queried for all inpatient and outpatient visits occurring from 1998 through 2018 by individuals who met the inclusion criteria of: Three or more total visits recorded in the EHR, 30 days or more between the first and last visits, and the existence of at least one encounter after age 10 and before age 90. For each patient, we analyzed all demographic, diagnostic, procedure, laboratory, and medication data recorded at each visit, as well the unstructured clinician notes.

### Natural Language Processing

In order to derive features from the unstructured clinician notes, we created a custom lexicon of suicide-relevant and psychiatric concepts using a variety of approaches including: (1) Selecting signs and symptoms, and mental and behavioral process semantic types from the Unified Medical Language System (UMLS)^11^; (2) Mapping DSM symptoms and concepts from structured instruments^12^; (3) Automatically extracting features from public sources including Wikipedia and MedScape; (4) Incorporating RDoC domain matrix terms^12^; (5) Selecting predictive features from coded suicide attempt prediction models^13^; and (6) Manual annotation of terms by expert clinicians. This lexicon was linked to UMLS concepts and included 480 distinct semantic concepts and 1,273 tokens or phrases. Using this lexicon, we ran the HiTex^14^ NLP named-entity extraction pipeline to identify concepts in over 120 million clinical notes. For each note, we identified the presence of a concept (e.g. symptom, disease, mental process) and further tagged concepts as negated (NEG), family history mention (FH) or negated family history (NFH). For negation and family history pipeline components, we utilized the ConText algorithm.^15^

### Case Definition

We have previously described the development of an EHR-based case definition for suicide.^9^ In summary, with the help of three expert clinicians, we identified codes from *International Classification of Diseases, Ninth Revision* (ICD-9) and *International Classification of Diseases, Tenth Revision* (ICD-10) that reliably captured suicide attempts with a positive predictive value (PPV) of greater than 0.70. Subjects having at least one of these codes were included in the case population. For cases, we also removed all data following the first suicide attempt (the index event), and made predictions at the penultimate visit prior to the index event. For the purpose of this study, the case definition was based solely on structured diagnostic information and did not include information derived from the clinician notes when classifying individuals as cases versus non-cases.

### Model Training

We split our data into training and testing sets with a 70/30 ratio, respectively. We applied two modeling approaches for suicide prediction. The first was a Naive Bayes Classifier (NBC) model, described in detail elsewhere.^16^ NBCs are a subclass of Bayesian networks that assume strong conditional independence of all input features, greatly reducing model complexity.^17^ NBCs have been shown to be well-suited for clinical decision support tasks and are highly scalable and interpretable; they compute a risk score for each concept using the odds ratios of its prevalence in case and non-case populations, ignoring interactions with other variables. During validation, the NBC risk scores for each concept in a patient’s visit history were added together to compute a cumulative suicide risk measure for the subject. If a patient had multiple instances of the same predictor over multiple visits, that predictor was counted multiple times at different visits of the patient. The NBC model was trained using *R* version 3.6.0 and the *R* packages *pROC* and *tidyverse*.

The second modeling approach was a Balanced Random Forest Classifier (BRFC),^18^ which unlike NBCs is capable of capturing interactions between features. Balanced Random Forests are an extension of Random Forest^19^ models, which work well with label-imbalanced datasets. Due to computational constraints, the BRFCs were trained and tested on a smaller subset of 140,000 subjects of the RPDR data. The occurrence rate of suicide attempts in our dataset is very low, at about 1%, resulting in low positive predictive values (PPV) on test sets with regular Random Forests. BRFCs balance the classes by either downsampling the majority class, upsampling the minority class, or resampling both classes with replacement during bootstrap draws until a specified ratio of classes is met. During the sampling of training data, we ensured that the proportion of cases was lifted from 1% to around 12%. The test set was left intact with the natural 1% suicide attempt rate. The data pipeline for arriving at training and testing sets for all described models is illustrated in Figure 2.

**Figure 2.**
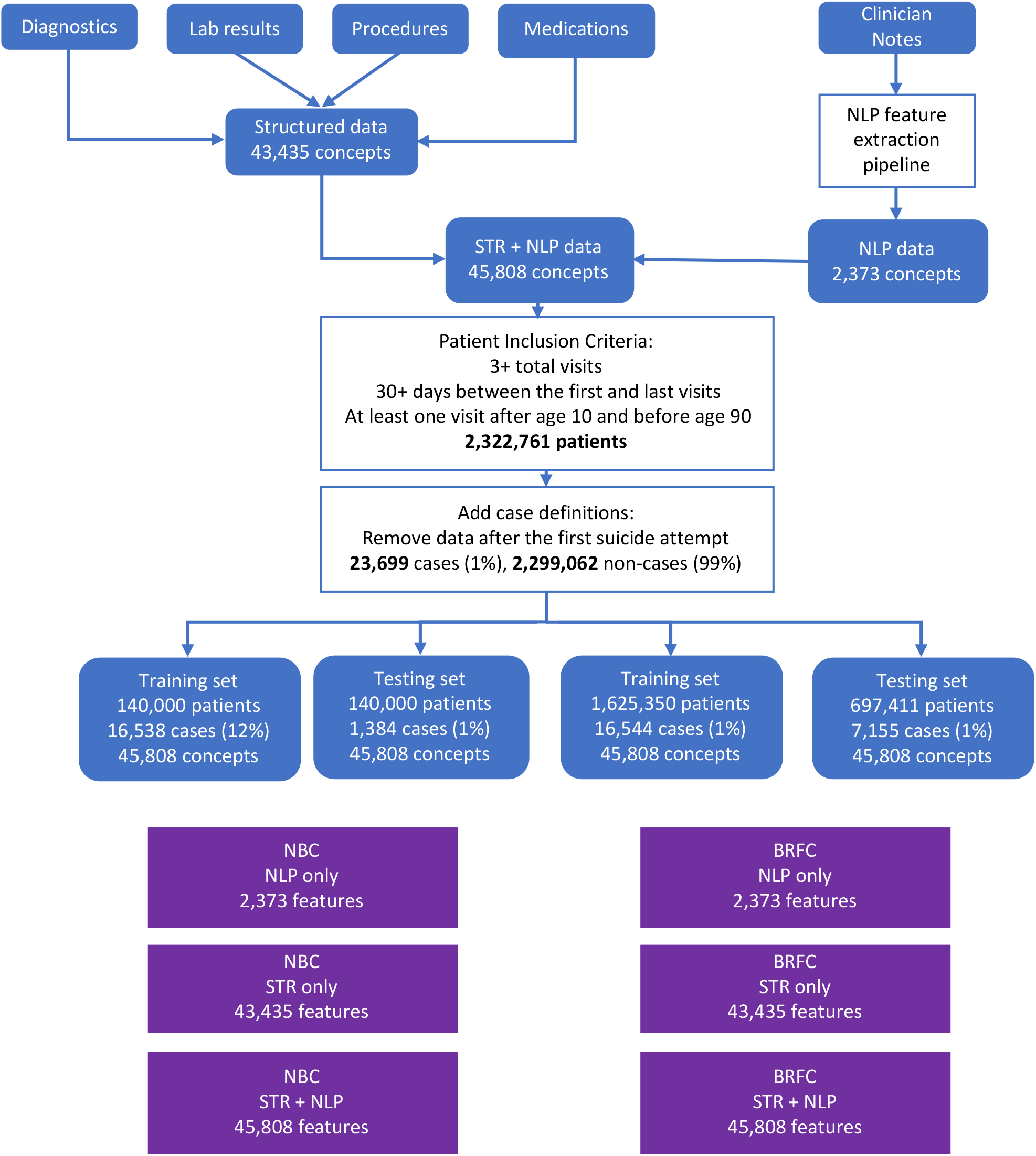
Data and Modeling Workflow. The diagram describes the filtering and processing steps taken to arrive at the final datasets used for training and testing different models described in this paper. STR – Structured Data; NLP – Unstructured data processed by Natural Language Processing; NBC – Naïve Bayesian Classifier; BRFC – Balanced Random Forest Classifier.

For selecting the parameters of the model, we performed a grid search with 5-fold cross-validation on the BRFC parameter space. Based on the grid search results, we arrived at a model with 30 trees, 50% of all features sampled for each tree, bootstrap sample size equal to the total number of samples, and 1:4 ratio of case to non-cases in every bootstrap sample, achieved with random undersampling of the majority class. Even after undersampling non-cases to 1:4 case:non-case ratio, the size of bootstrap samples remained sufficiently large due to the relatively high case prevalence (12%) in the training data. We used *Python* version 3.6.9 with the libraries *scikit-learn, imblearn, numpy, pandas*, and *matplotlib*. The packages *imblearn and scikit-learn* were useful for training and testing balanced random forests. Libraries *numpy* and *pandas* were helpful for data transformations and analyses. Paper visualizations were produced using *matplotlib*.

We used area under the receiver operating characteristic curve (AUC) as the primary predictive performance metric. In order to create confidence intervals and enable comparison of AUC values of different models, we used the percentile bootstrapping method with a simulation size of 1,000. We also measured PPV and sensitivity over a range of specificities. Since the primary goal of our work was to investigate properties of the NLP dataset rather than to build an optimal predictive model, we maximized simplicity in the study design: All predictions were made at the visit prior to the first suicide attempt for cases, and the last visit recorded for non-cases.

### Contingency Analysis

In order to better understand the interactions between structured and unstructured data, we performed a separate contingency analysis to identify interactions between structured and unstructured features that differed substantially between case and non-case populations. To account for possible effects of sample size differences between case and non-case populations, we randomly sampled two equal cohorts --one with 23,566 cases and the other with 23,566 non-cases. (These cohorts were sampled from the original dataset before training and testing splits were made.) To simplify analysis, we counted each feature only at its first occurrence for each subject.

For simplicity in the following discussion, we will refer to a feature derived from structured data as *A*, and a feature derived from natural language processing of unstructured data as *B*. For each feature pair *A-B*, we computed contingency tables for both case and non-case populations (Table 1). To measure the strength of association between feature *A* and feature *B* within each cohort, we performed a Chi-squared test of independence. The null hypothesis was that *A* is independent of *B*, while the alternative hypothesis was that there is an association between *A* and *B*. We computed the statistic *T*_*i*_ for both case and non-case populations:

**Table 1.** Contingency tables of structured-unstructured concept pairs *A-B*, for case and non-case cohorts.

| Cases |  |  | Non-Cases |  |  |
| --- | --- | --- | --- | --- | --- |
| Concept | B: 1 | B: 0 | Concept | B: 1 | B: 0 |
| A: 1 | $a_1$ | $b_1$ | A: 1 | $a_0$ | $b_0$ |
| A: 0 | $c_1$ | $d_1$ | A: 0 | $c_0$ | $d_0$ |

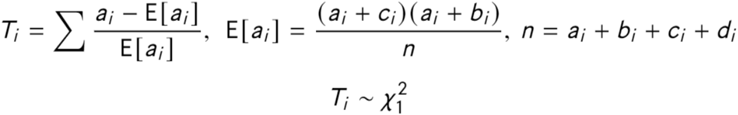

where *a, b, c*, and *d* are as defined in Table 1. Under the null hypothesis, *T*_*i*_ follows a Chi-squared distribution with one degree of freedom. This value can be used to compute p-values from the Chi-squared quantile function.

In order to determine whether the interactions between feature *A* and feature *B* differed between case and non-case populations, we used Woolf’s method for testing for homogeneity.^*20*^ The null hypothesis was that the odds ratios computed on each of the case and non-case populations were equal, while the alternative hypothesis was that these differed significantly. We calculated Woolf’s test statistic (*X*^2^_HOM_) as follows:

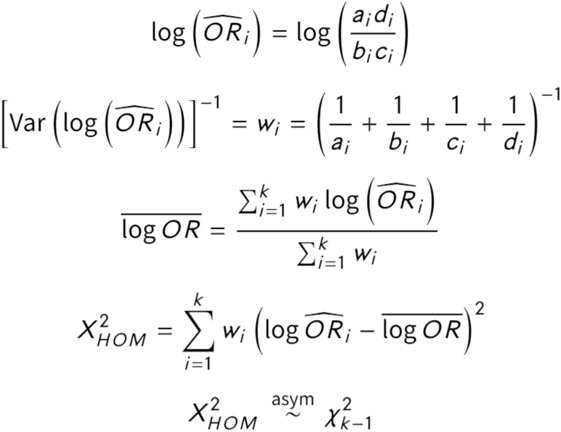

For *k = 0*, under the null hypothesis, *X*^2^_HOM_ follows a Chi-squared distribution with one degree of freedom. For clarity, we will refer to Woolf’s test statistic *X*^2^_HOM_ as *Interaction Heterogeneity* (IH). Interaction heterogeneity provides a summary measure of the difference in the overall shape of the contingency table between case and non-case populations.

Next, we examined the joint distribution p(*AB*|*Y*), conditional on the case variable *Y* (suicide vs. non-suicide). Using Bayes’ rule, this distribution can be used to derive the more clinically interesting distribution p(*Y*|*AB*) --specifically P(*Y* = 1 | *A* = 1, *B* = 1) --which is the probability of the patient attempting suicide in the future given that the patient has both features *A* and *B*:

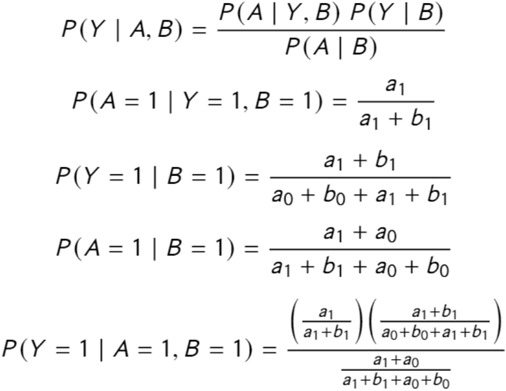

The variables *a*_*i*_, *b*_*i*_, *c*_*i*_, *d*_*i*_ are as in Table 1, except that the entries in the contingency table of cases have been divided by 100 to reflect the 1/99 case-non-case ratio encountered in the clinical population. Thus, using Woolf’s method, we are able to identify specific structured-unstructured feature interactions that are most different between case and non-case cohorts, and thus have the most potential for improving predictive performance.

Combining the above methods, we assembled a list of structured-unstructured feature pairs *AB* in which: 1) Both *A* and *B* were among the top 200 most important features as ranked by the absolute value of the NBC feature risk scores; 2) The joint occurrence of *A* and *B* were significantly different from the expected value under the null within both case and non-case cohorts, as measured using the Chi-squared statistic *T*_*i*_; and 3) The interaction between *A* and *B* was significantly different (heterogeneous) between the case population and the non-case population --as measured by interaction heterogeneity (IH). For ease of interpretation, we included only unstructured features that were either “positive” or “positive family history” mentions, and excluded “negative” and “negative family history” mentions.

Since the goal of this analysis was not to simply find meaningful interactions in the dataset, but rather to identify meaningful interactions between structured and unstructured features, we performed the contingency analysis on structured-unstructured feature pairs, but not on structured-structured or unstructured-unstructured feature pairs.

### Ethics

This research was approved by the Mass General Brigham Institutional Review Board, along with an IRB reliance agreement from the Boston Children’s Hospital Institutional Review Board.

## Results

Applying the inclusion and exclusion criteria to the extracted data yielded 1,625,350 training subjects for the NBC models, which included 1,608,806 non-cases (99%) and 16,544 cases (1%) (Figure 2). The testing set consisted of 697,411 subjects, including 7,155 cases. For the BRFC models, the dataset included 140,000 subjects for each of the training and testing populations, with the former having 16,538 cases (12%, due to the sampling approach mentioned above) and the latter having 1,384 (1%, reflecting the prevalence in the clinical population). For both experiments, we had the same set of 45,808 features which included 43,435 structured features (95%) and 2,373 features derived from unstructured data using NLP (5%).

### Model Performance

The results of training and testing are presented in Table 2. We found that for both NBC and BRFC modelling approaches, training on structured data features resulted in higher predictive performance than training on features derived from unstructured data, with an improvement in AUC of 2-3% (p < 0.001).

**Table 2a.**
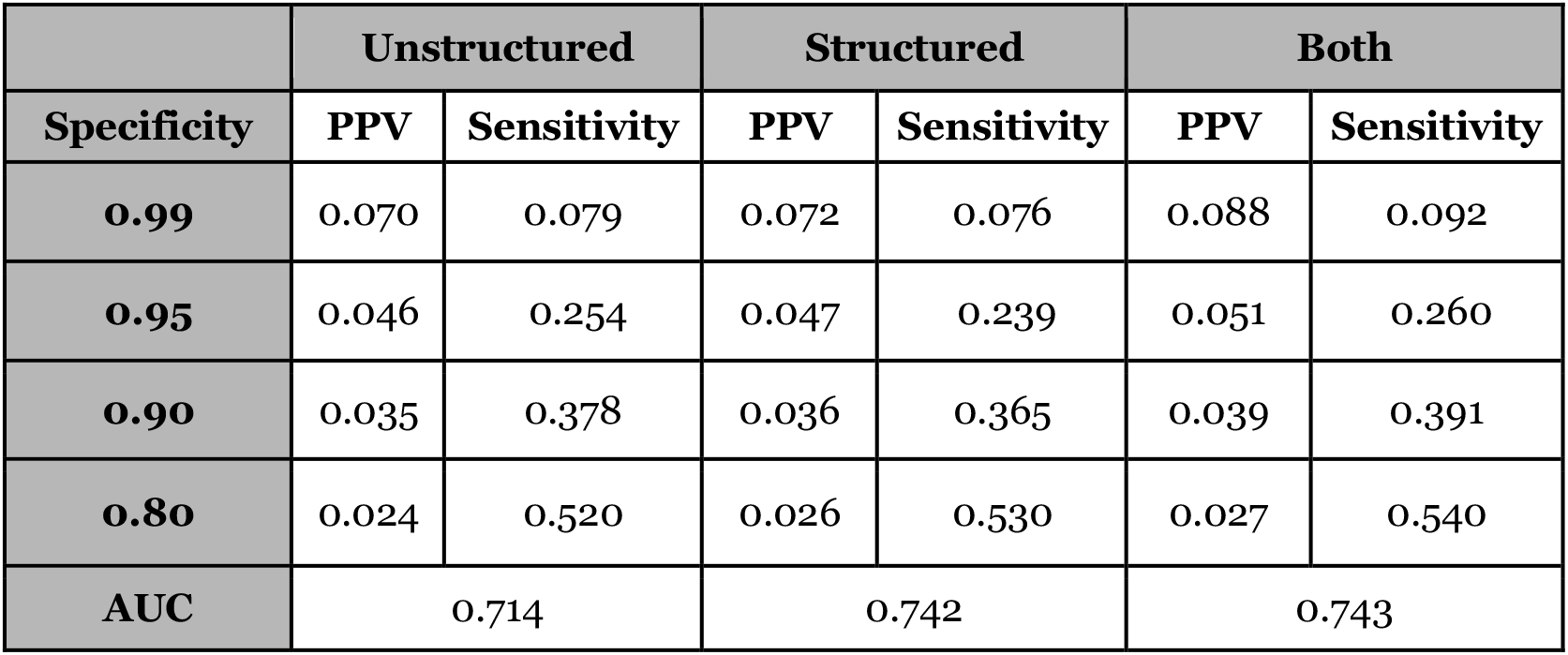
Performance of NBC models on the test set. There is no significant increase (p = 0.688) in AUC between the model based on structured-data-only and the model based on both structured and unstructured data.

**Table 2b.** Performance of BRF models on the test set. There is a significant increase (p < 0.001) in AUC between the model based on structured-data-only and the model based on both structured and unstructured data.

|  | Unstructured |  | Structured |  | Both |  |
| --- | --- | --- | --- | --- | --- | --- |
| Specificity | PPV | Sensitivity | PPV | Sensitivity | PPV | Sensitivity |
| **0.99** | 0.142 | 0.168 | 0.191 | 0.246 | 0.219 | 0.267 |
| **0.95** | 0.082 | 0.447 | 0.092 | 0.507 | 0.097 | 0.545 |
| **0.90** | 0.057 | 0.608 | 0.063 | 0.657 | 0.066 | 0.697 |
| **0.80** | 0.037 | 0.766 | 0.040 | 0.820 | 0.041 | 0.845 |
| **AUC** | 0.868 |  | 0.887 |  | 0.902 |  |

For the NBC model, training on *both* structured and unstructured data yielded no significant improvement over training on structured data alone (p-value = 0.67). However, for BRFCs, training with both structured and unstructured data led to a moderate but significant 1.6% increase in AUC relative to training on structured data alone (p-value <0.001). The combined structured and unstructured BRFC model also exhibited moderate increases in PPV and sensitivity from the structured-data-only model across all specificity thresholds, with a 4% increase in sensitivity at both 0.90 and 0.95 specificity, in addition to increases in PPV.

### Contingency Analysis

Table 3 shows structured-unstructured feature pairs in which the relationship between the two features differed most between case and non-case cohorts --namely, those with the highest interaction heterogeneity. Table 3a shows feature pairs in which the structured feature *A* was associated with greater suicide risk (i.e. feature *A* occurred more frequently in the case cohort than in the non-case cohort). These include drug and opioid use, suicidal ideation, and borderline personality disorder which are associated with various high-risk NLP features including schizophrenia, self-reported suicide attempts, imprisonment, and homelessness.

**Table 3a.** Structured-unstructured feature pairs *AB* with high interaction heterogeneity (IH), where *A* is a strong *risk factor* for suicide attempt. A high IH value indicates that the relationship between A and B changes significantly between case and non-case populations.

| Features |  | Cases |  |  |  | Non-Cases |  |  |  |  |
| --- | --- | --- | --- | --- | --- | --- | --- | --- | --- | --- |
| Structured (A) | Unstructured (B) | A | B | AB Expected | AB Actual | A | B | AB Expected | AB Actual | IH |
| Other, mixed, or unsp. Drug abuse, unsp. Use | suicide attempts | 2356 | 3741 | 374.53 | 1003 | 148 | 563 | 3.54 | 53 | 77.55 |
| Other, mixed, or unsp. Drug abuse, unsp. Use | section XII | 2356 | 3045 | 304.85 | 849 | 148 | 403 | 2.53 | 43 | 74.72 |
| Other, mixed, or unsp. Drug abuse, unsp. Use | living on the street | 2356 | 1113 | 111.43 | 532 | 148 | 154 | 0.97 | 36 | 66.66 |
| Other, mixed, or unsp. Drug abuse, unsp. Use | prison | 2356 | 2043 | 204.53 | 825 | 148 | 358 | 2.25 | 51 | 62.57 |
| Other, mixed, or unsp. Drug abuse, unsp. Use | intoxications | 2356 | 2663 | 266.61 | 889 | 148 | 462 | 2.91 | 50 | 60.56 |
| Suicidal ideation | section XII | 1820 | 3045 | 235.49 | 1299 | 127 | 403 | 2.17 | 81 | 54.69 |
| Other, mixed, or unsp. Drug abuse, unsp. Use | undomiciled | 2356 | 2357 | 235.97 | 964 | 148 | 408 | 2.57 | 55 | 54.50 |
| Other, mixed, or unsp. Drug abuse, unsp. Use | opioid dependence | 2356 | 1625 | 162.69 | 841 | 148 | 195 | 1.23 | 44 | 53.86 |
| Suicidal ideation | schizoaffective schizophrenia | 1820 | 676 | 52.28 | 223 | 127 | 118 | 0.64 | 21 | 52.75 |
| Other, mixed, or unsp. Drug abuse, unsp. Use | sober | 2356 | 3667 | 367.12 | 1329 | 148 | 723 | 4.55 | 76 | 52.29 |
| Other, mixed, or unsp. Drug abuse, unsp. Use | unspecified bipolar disorder | 2356 | 3488 | 349.20 | 932 | 148 | 699 | 4.40 | 49 | 48.53 |
| Other, mixed, or unsp. Drug abuse, unsp. Use | schizoaffective schizophrenia | 2356 | 676 | 67.68 | 172 | 148 | 118 | 0.74 | 15 | 46.44 |
| Opioid abuse, unspec. Use | sober | 1305 | 3667 | 203.35 | 710 | 78 | 723 | 2.40 | 42 | 46.09 |
| Other, mixed, or unsp. Drug abuse, unsp. Use | methadone | 2356 | 2992 | 299.54 | 1165 | 148 | 653 | 4.11 | 69 | 45.55 |
| Borderline personality | methadone | 582 | 2992 | 74.00 | 139 | 35 | 653 | 0.97 | 14 | 43.59 |
| Opioid abuse, unspec. Use | living on the street | 1305 | 1113 | 61.72 | 293 | 78 | 154 | 0.51 | 18 | 43.28 |
| Opioid type dependence, continuous use | drug seeking | 710 | 463 | 13.97 | 96 | 50 | 51 | 0.11 | 9 | 37.61 |
| Suicidal ideation | suicidality | 1820 | 2546 | 196.90 | 1057 | 127 | 380 | 2.05 | 58 | 35.84 |
| Other, mixed, or unsp. Drug abuse, unsp. Use | cluster b | 2356 | 495 | 49.56 | 175 | 148 | 43 | 0.27 | 10 | 35.70 |
| Unspec. Neurotic disorder | opioid dependence | 1003 | 1625 | 69.26 | 191 | 72 | 195 | 0.60 | 12 | 35.48 |

Table 3b shows feature pairs in which the structured feature *A* was associated with lower suicide risk (i.e. *A* occurred less frequently in the case cohort than in the non-case cohort). These include concepts such as annual exams, mammograms, and tumor screenings that are associated with NLP concepts such as impulse-control disorder and use of hallucinogenic and psychoactive drugs derived from psilocybin mushrooms (referred to as “vacuuming” in informal parlance). In many cases, structured codes such as mammograms and tumor screenings are confounded with older age which is protective of suicide. Hence lower suicide risk associated with interaction of these structured variables with high-risk concepts such as impulse-control disorder and hallucinogenic drug use is to be expected. (In Table 3, “AB Expected” corresponds to E[a_i_] used in computation of the T_i_ statistic defined above.)

**Table 3b.** Structured-unstructured feature pairs *AB* with high interaction heterogeneity (IH), where *A* is a strong *protective factor* against suicide. A high IH value indicates that the relationship between A and B changes significantly between case and non-case populations. Among the unstructured concepts, “ICD” refers to impulse-control disorder, and “vacuuming” refers to use of hallucinogenic and psychoactive drugs derived from psilocybin mushrooms.

| Features |  | Cases |  |  |  | Non-Cases |  |  |  |  |
| --- | --- | --- | --- | --- | --- | --- | --- | --- | --- | --- |
| Structured (A) | Unstructured (B) | A | B | AB Expected | AB Actual | A | B | AB Expected | AB Actual | IH |
| Screening mammogram for malignant neoplasm of breast | impulse-control disorder (ICD) | 89 | 661 | 2.50 | 51 | 2091 | 3658 | 325.03 | 875 | 110.08 |
| Annual Exam | ICD | 171 | 661 | 4.80 | 81 | 2596 | 3658 | 403.53 | 1249 | 94.20 |
| Screening mammogram for malignant neoplasm of breast | vacuuming | 89 | 231 | 0.87 | 25 | 2091 | 1546 | 137.37 | 374 | 93.77 |
| Screening digital breast tomosynthesis, bilateral | ICD | 103 | 661 | 2.89 | 46 | 1656 | 3658 | 257.41 | 730 | 71.63 |
| Encounter for screening, unspec. | ICD | 55 | 661 | 1.54 | 30 | 809 | 3658 | 125.75 | 344 | 66.36 |
| Screening digital breast tomosynthesis, bilateral | vacuuming | 103 | 231 | 1.01 | 23 | 1656 | 1546 | 108.79 | 332 | 62.36 |
| Encounter for screening for malignant neoplasm of colon | ICD | 61 | 661 | 1.71 | 31 | 1399 | 3658 | 217.46 | 620 | 57.69 |
| Screening mammogram for malignant neoplasm of breast | ICD | 89 | 2019 | 7.64 | 80 | 2091 | 10987 | 976.24 | 1765 | 53.97 |
| Pure hypercholesterolemia, unsp. | ICD | 64 | 661 | 1.80 | 30 | 1328 | 3658 | 206.43 | 596 | 49.89 |
| Screening digital breast tomosynthesis, bilateral | ICD | 103 | 2019 | 8.84 | 82 | 1656 | 10987 | 773.15 | 1422 | 44.84 |
| Annual Exam | vacuuming | 171 | 231 | 1.68 | 23 | 2596 | 1546 | 170.54 | 423 | 44.53 |
| Physical therapy evaluation low complex 20 mins | ICD | 36 | 661 | 1.01 | 22 | 678 | 3658 | 105.39 | 325 | 44.29 |
| Screening, malig. neopl. colon | vacuuming | 61 | 231 | 0.60 | 14 | 1399 | 1546 | 91.91 | 269 | 43.32 |
| Screening, malig. neopl. breast | ICD | 30 | 661 | 0.84 | 18 | 571 | 3658 | 88.76 | 272 | 36.53 |
| Other hemorrhoids | ICD | 37 | 661 | 1.04 | 17 | 559 | 3658 | 86.89 | 236 | 33.29 |
| Age-related osteoporosis without current pathological fracture | ICD | 32 | 661 | 0.90 | 18 | 549 | 3658 | 85.34 | 271 | 32.33 |
| Asymptomatic menopausal state | vacuuming | 20 | 231 | 0.20 | 7 | 387 | 1546 | 25.42 | 81 | 29.70 |
| Other melanin hyperpigmentation | vacuuming | 25 | 231 | 0.25 | 8 | 699 | 1546 | 45.92 | 156 | 29.59 |
| Screening, unspecified | ICD | 55 | 2019 | 4.72 | 46 | 809 | 10987 | 377.70 | 692 | 29.58 |
| Mod sed same phys/qhp each addl 15 mins | ICD | 28 | 661 | 0.79 | 13 | 822 | 3658 | 127.77 | 329 | 28.45 |

As described above, interaction heterogeneity (IH) provides a summary measure of the difference in the overall shape of the contingency tables between case and non-case populations. In order to provide a more intuitive understanding of IH, Tables 4a and 4b provide illustrative examples of contingency tables for two structured-unstructured feature pairs *AB*: One with a high IH value of 77.55 (“Other, mixed, or unspecified drug abuse, unspecified use” & “suicide attempts”), and the other with a low IH value of 3.95 (“Opioid abuse, unspecified use” & “junk (heroin)”). For simplicity, we refer to the number of individuals who had both *A* and *B* in the cases cohort as *AB*_*cases*_, and to the number of people who had *A* but did not have *B* in the cases cohort as A∼B_cases_, and so forth.

**Table 4a.** Contingency tables for the structured-unstructured pair “Other, mixed, or unspecified drug abuse, unspecified use” (*A*) and “suicide attempts” (*B*). This feature pair has a high interaction heterogeneity (IH) value of 77.55. Values shown are proportions of the total number of samples (23,566) for each bin.

| **Cases** |  |  | **Non-Cases** |  |  |
| --- | --- | --- | --- | --- | --- |
| **Concept** | **B: 1** | **B: 0** | **Concept** | **B: 1** | **B: 0** |
| **A: 1** | 0.0401 | 0.0541 | **A: 1** | 0.0021 | 0.004 |
| **A: 0** | 0.1095 | 0.7376 | **A: 0** | 0.0204 | 0.9150 |

**Table 4b.** Contingency tables for the structured-unstructured pair “Opioid abuse, unspecified use” (*A*) and “junk (heroin)” (*B*). This feature pair has a low IH value of 3.95. Values shown are proportions of the total number of samples (23,566) for each bin. The differences between the two distributions are smaller in Table 4b than in Table 4a, resulting in a lower IH value.

| *Cases* |  |  | *Non-Cases* |  |  |
| --- | --- | --- | --- | --- | --- |
| **Concept** | **B: 1** | **B: 0** | **Concept** | **B: 1** | **B: 0** |
| **A: 1** | 0.0443 | 0.0079 | **A: 1** | 0.0022 | 0.0010 |
| **A: 0** | 0.1071 | 0.7820 | **A: 0** | 0.0297 | 0.9085 |

The values for AB_cases_ and AB_non-cases_ are similar for both pairs of contingency tables (Tables 4a and 4b), as are the values for ∼AB_cases_ and ∼AB_non-cases_. However, the differences between A∼B_cases_ and A∼B_non-cases_, and the differences between ∼A∼B_cases_ and ∼A∼B_non-cases_ are greater in Table 4a than in Table 4b. Thus, the overall shape of the contingency table in Table 4a changes more between case and non-case populations than the contingency in Table 4b. This yields a larger IH value for Table 4a and a smaller IH value for Table 4b, indicating that the interaction of concepts in Table 4a is more strongly associated with the suicide-attempt outcome.

In order to study the difference between IH and more traditional measures of risk, Figure 4 plots IH versus the *joint suicide attempt risk* of features *A* and *B* (defined as the log of the ratio of the expected joint occurrences of *AB* in the case vs. non-case cohorts). As mentioned, IH is a measure of whether the *interaction* between features *A* and *B* differs significantly between case and non-case cohorts. The *joint suicide attempt risk* provides a summary measure of association between the features and the outcome, reflecting the difference in the number of occurrences of *A* and *B* between case and non-case cohorts. (To reduce noise, we only included feature pairs *AB* with at least 10 joint occurrences in either case or non-case cohorts.)

**Figure 3.**
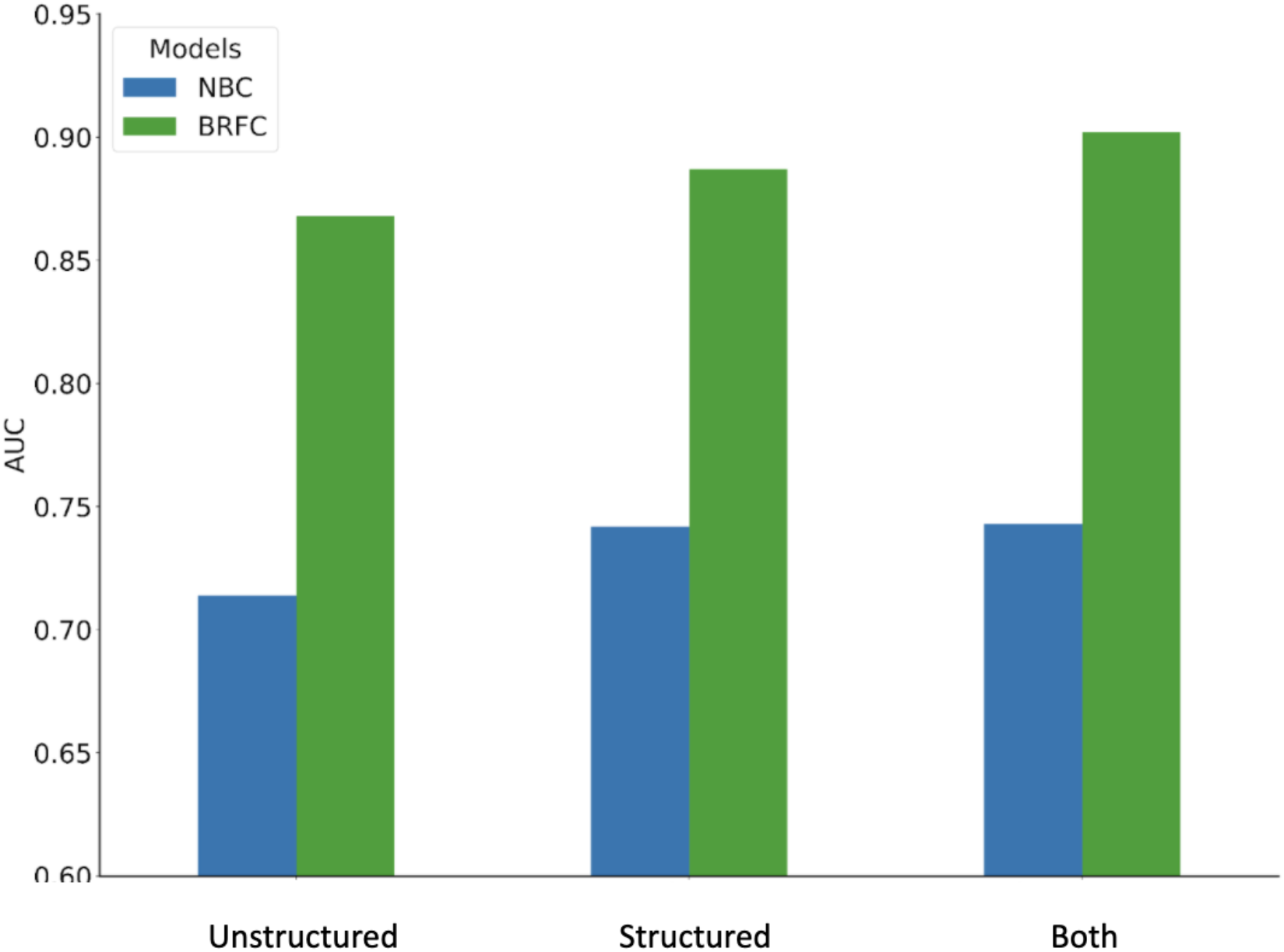
Performance of NBC and BRFC models, by type of data used. BRFC models perform considerably better than NBC models in terms of AUC across all three datasets. Combining structured and unstructured data yields better performance than using structured data alone, which itself performs better than using unstructured data only.

**Figure 4.**
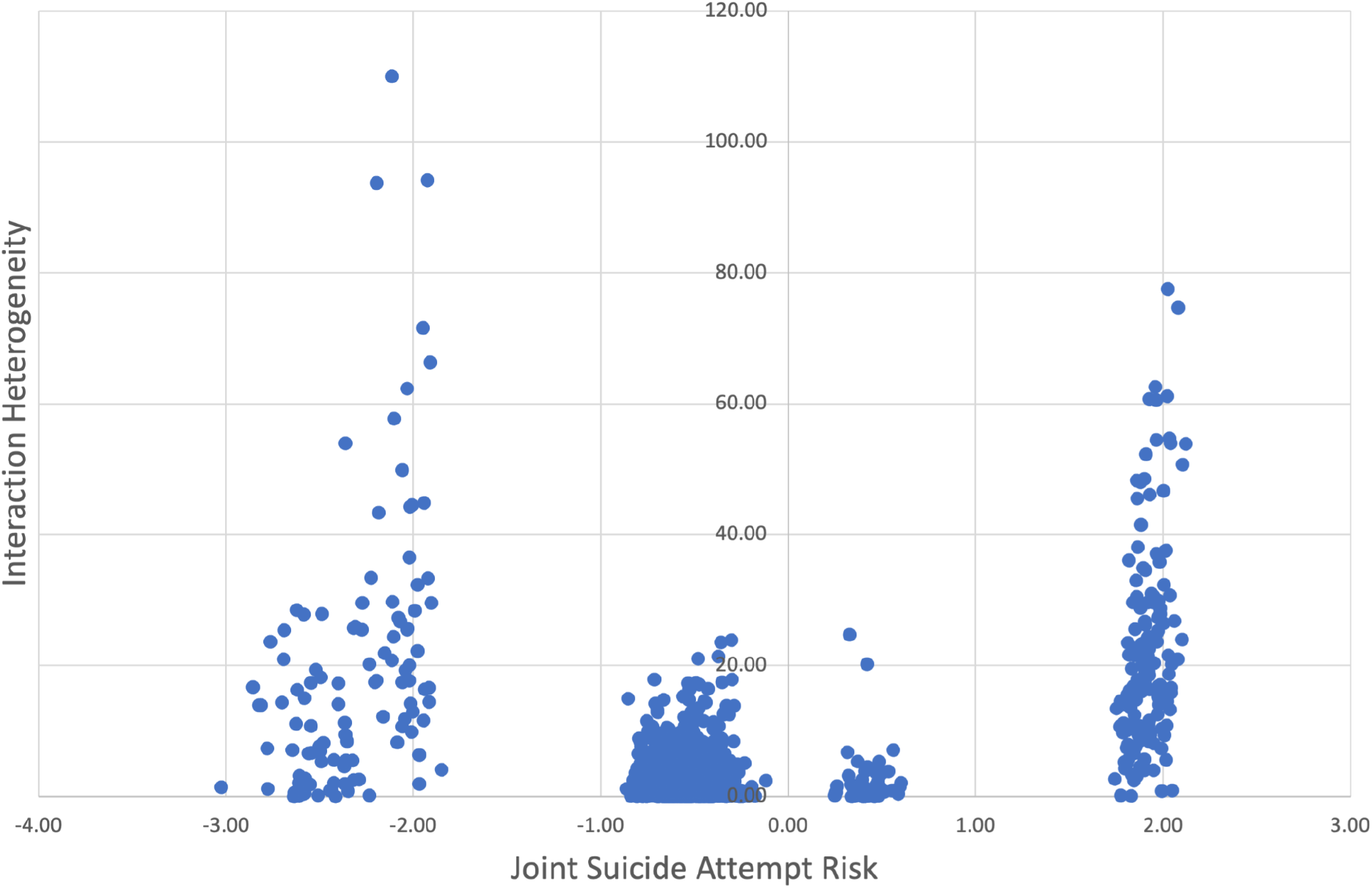
Interaction heterogeneity versus joint suicide risk. A comparison of joint suicide attempt risk and interaction heterogeneity. Each data point corresponds to a structured-unstructured feature pair *AB*. The x-axis shows the joint suicide risk of features *A* and *B*, defined as the log of the ratio of the expected joint occurrences of *AB* in the case vs. non case cohorts. The y-axis shows the interaction heterogeneity, a measure of how much the interaction between *A* and *B* differs between case and non-case cohorts. The plot shows that feature pairs with similar joint suicide attempt risk can have very different interaction heterogeneity.

Figure 4 shows that many feature pairs with similar joint suicide risk have a large variation in IH --highlighting the fact that IH can reveal variation in feature interactions that the ratio of expected occurrences does not capture.

This is illustrated further in Table 5, which presents interactions that correspond to the rightmost cluster in Figure 4 (i.e. feature pairs with joint suicide risk between 1.7 and 2.3). Within this cluster, Table 5a presents the 20 feature interactions with the highest values of IH, and Table 5b presents the 20 feature interactions with the lowest values of IH. Although the joint suicide risk values are approximately the same in both tables, we see that the nature of interactions is different between Tables 5a and 5b. Table 5a contains mostly general substance-abuse structured features (e.g. “Other, mixed, or unspecified drug abuse, unspecified use”), while Table 5b includes specific substance-abuse structured features such as cocaine, methadone, barbiturate, and opioid consumption. Furthermore, the substance abuse codes in Table 5a interact mostly with non-substance-abuse unstructured features such as “lack of domicile”, “schizophrenia” and “imprisonment”, while the substance-abuse codes in Table 5b interact mostly with other substance-abuse-related unstructured features -most prominently, heroine and thioridazine. Thus, interactions between features that are near-synonyms show less difference between case and non-case cohorts than interactions between features that are more heterogeneous.

**Table 5a.** Structured-unstructured feature pairs *A-B* with high interaction heterogeneity (IH) values. The joint suicide attempt risk of features *A* and *B* is defined as the log of the ratio of the expected joint occurrences of *AB* in the case vs. non case cohorts.

| Structured Feature (*A*) | Unstructured Feature (*B*) | Joint Suicide Attempt Risk | IH |
| --- | --- | --- | --- |
| Other, mixed, or unspecified drug abuse, unspecified use | suicide attempts | 2.02 | 77.55 |
| Other, mixed, or unspecified drug abuse, unspecified use | section XII | 2.08 | 74.72 |
| Other, mixed, or unspecified drug abuse, unspecified use | living on the street | 2.06 | 66.66 |
| Other, mixed, or unspecified drug abuse, unspecified use | prison | 1.96 | 62.57 |
| Other, mixed, or unspecified drug abuse, unspecified use | undomiciled | 2.02 | 61.18 |
| Other, mixed, or unspecified drug abuse, unspecified use | intoxications | 1.96 | 60.56 |
| Suicidal ideation | section XII | 2.03 | 54.69 |
| Other, mixed, or unspecified drug abuse, unspecified use | undomiciled | 1.96 | 54.50 |
| Other, mixed, or unspecified drug abuse, unspecified use | opioid dependence | 2.12 | 53.86 |
| Suicidal ideation | schizoaffective schizophrenia | 1.91 | 52.75 |
| Other, mixed, or unspecified drug abuse, unspecified use | sober | 1.91 | 52.29 |
| Opioid abuse, unspecified use | methadone | 2.02 | 48.85 |
| Other, mixed, or unspecified drug abuse, unspecified use | unspecified bipolar disorder | 1.90 | 48.53 |
| Suicidal ideation | delusions | 1.86 | 48.32 |
| Other, mixed, or unspecified drug abuse, unspecified use | methadone | 2.00 | 46.72 |
| Other, mixed, or unspecified drug abuse, unspecified use | schizoaffective schizophrenia | 1.96 | 46.44 |
| Opioid abuse, unspecified use | sober | 1.93 | 46.09 |
| Other, mixed, or unspecified drug abuse, unspecified use | methadone | 1.86 | 45.55 |
| Cocaine abuse, unspecified use | methadone | 1.97 | 43.78 |
| Borderline personality | methadone | 1.88 | 43.59 |

**Table 5b.**
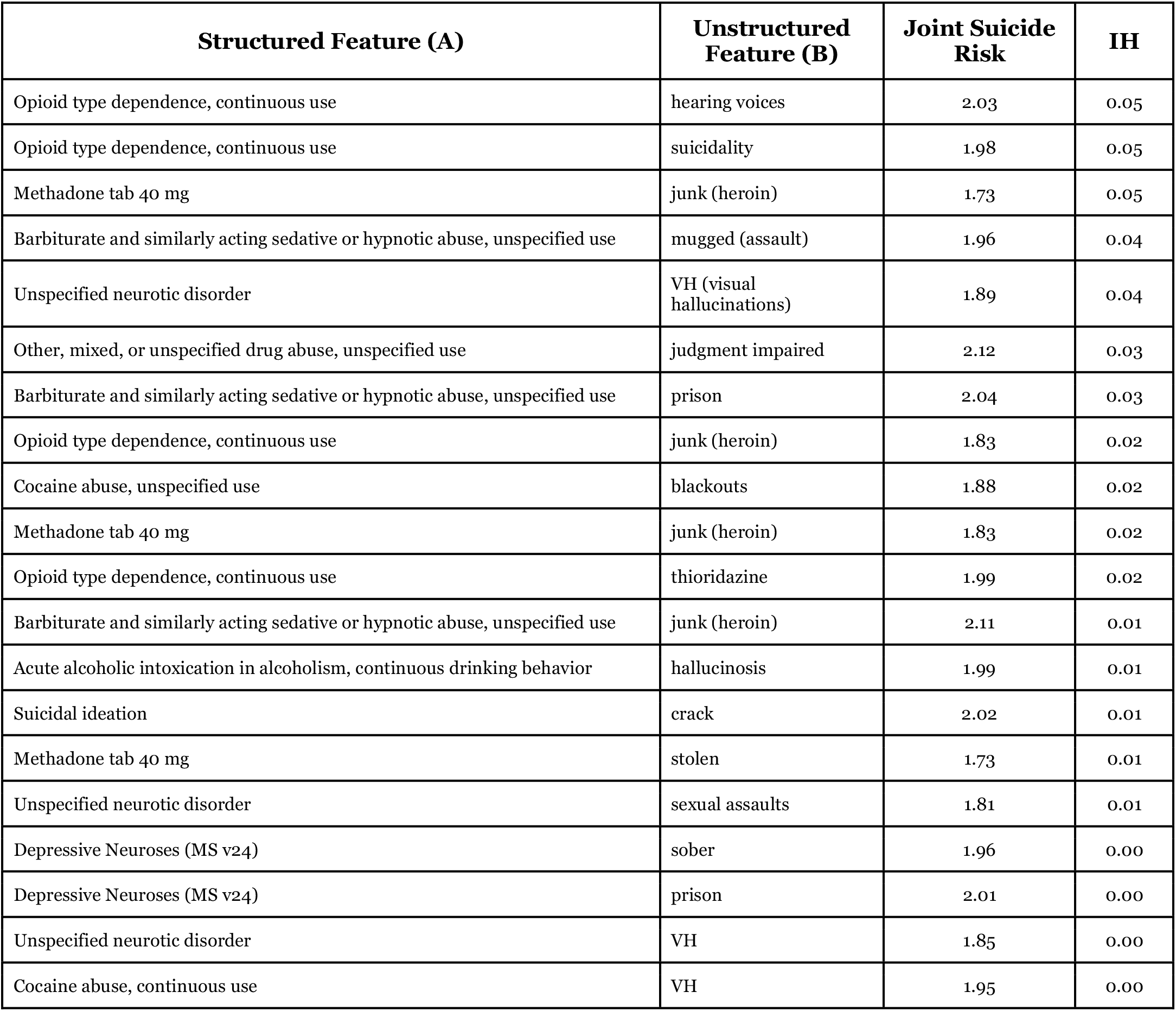
Structured-unstructured feature pairs *A-B* with low interaction heterogeneity (IH) values. The joint suicide attempt risk of features *A* and *B* is defined as the log of the ratio of the expected joint occurrences of *AB* in the case vs. non case cohorts.

## Discussion

We found that models trained only on features derived from structured-data perform better than models trained only on features derived from unstructured data. The performance gap between models trained with structured data and those trained with unstructured data is quite small, considering the compact size of the unstructured data.

Combining unstructured data with structured data provided almost no performance benefit with the NBC model, whereas the BRFC model showed a significant increase in AUC. The fact that the NBC model only negligibly benefitted from the addition of NLP concepts is not surprising; while interactions between structured and unstructured features could contain useful signals, NBCs assume conditional independence among features, and so cannot exploit these interactions to improve predictive performance. On the other hand, BRFCs are designed to capture interactions between features, and are thus able to deliver a significant improvement in predictive performance. Indeed, examining trees in the BRFC model, we found many examples where splits based on NLP concepts were either preceded or followed by structured-data-based splits, bearing evidence that the BRFC models captured useful structured-unstructured interactions.

Structured-unstructured feature pairs whose interactions differed most between suicidal and non-suicidal populations were those that described heterogeneous pairs of general concepts, rather than pairs of similar concepts. Such insights into the changing nature of feature interactions between case and non-case cohorts can help to improve predictive performance and provide a deeper understanding of clinical risk.

This study is subject to a number of limitations. We analyzed 20 years of longitudinal healthcare data from a single healthcare system including hospital admissions, observational stays, emergency department visits and outpatient encounters. Visits outside this geographical setting, time period, and network of hospitals were not included, and therefore this study dataset may be missing some encounters which could have potentially been useful for predicting suicide attempts. Moreover, some of these excluded visits may have been for suicidal behavior, meaning that some patients may have been incorrectly identified as non-case subjects or correctly identified as case subjects but given incorrect onset times. For patient diagnoses, we included both ICD-9 and ICD-10 codes since both encoding standards were used in the RPDR during the last 20 years. Due to this, there are somee concepts for which both ICD-9 and ICD-10 definitions have been included in the dataset, adding extra computational burden. Since the goal of this research was to investigate properties of structured and unstructured data, we compared predictive performance of NBCs and BRFCs, which are relatively easy to interpret. To achieve a potentially superior predictive model, it would also be worthwhile to consider other modelling approaches such as XGBoost, neural networks, and support vector machines, as well as complex feature selection techniques such as PCA and t-SNE. However, these modelling methods are more difficult to interpret, making them less suitable for the present study. They are potential avenues for future work.

Another limitation is that suicide attempt risk predictions were performed only on the penultimate visits prior to a suicide attempt. This was done to reduce the complexity and computational burden of the prediction task while allowing us to focus on differences between structured and unstructured features. As a result, the specific models developed here are designed to predicting risk in later visits of patients and may not predict suicide risk sufficiently in advance if used in earlier visits. Predictive models trained for practical purposes would be designed for predicting at any point during the patient’s longitudinal history. One approach for doing this with random forests is to sample random visits in the patient’s medical timeline and include cumulative feature history up until that visit as “snapshots.” We have explored such multi-temporal suicide risk predictions with random forests in a separate study.^21^

Previous studies have examined the use of unstructured EHR data in clinical prediction models in general, and in suicide prediction models in particular. Tsui et al.^1^ showed that the use of NLP features extracted from clinician notes significantly improved the AUC of an ensemble of extreme gradient boosting models and of a Lasso model over a structured-data only baseline model. Poulin et al. used keywords extracted from unstructured clinician notes to predict suicide risk among US veterans with an accuracy of 65%.^4^ Carson et al constructed a random forest model trained on structured and unstructured EHR data of psychiatrically hospitalized adolescents to predict suicidal behavior with an AUC of 0.68.^22^

In the present study, we examined the integration of features derived from unstructured clinician notes into structured-data-based suicide risk prediction models. We showed that a model that assumes independence among variables (NBC) does not significantly benefit from addition of unstructured features, whereas models such as Balanced Random Forest Classifiers that explicitly capture interactions exhibit performance increases when unstructured features are added. We also proposed and implemented a framework for identifying specific structured-unstructured feature pairs whose interaction patterns differ with respect to a patient’s suicide risk, and thus have the potential to improve predictive performance and increase understanding of clinical risk. These findings and this framework can be used to improve current and future EHR-based clinical prediction models, which are becoming increasingly widespread in clinical settings.

## Data Availability

The data used in this study cannot be made publicly available due to restrictions relating to the use of electronic health record data.

## Acknowledgements

This work was supported in part by a gift from the Tommy Fuss Fund and R01MH117599 (Drs. Smoller and Reis) from the National Institute of Mental Health.

## Author Contributions

IB and BYR conceived and planned the research, together with input from all other authors. IB conducted the principal data analysis and modeling work. VC conducted the NLP analysis and prepared the datasets for analysis. YBC also contributed to the modeling work. BYR supervised the research. IB, VC, YBC, EM, MKN, JWS and BYR all discussed the results and contributed to the final manuscript.

